# Design and evaluation of an automated real-time SOFA score calculator in an electronic health record system, for early sepsis diagnosis

**DOI:** 10.1101/2024.09.30.24314269

**Authors:** Gustaf Smitt Einarson, Hampus Nordqvist, Ulrika Marking, Sandra Jonmarker, Åsa Parke, Christian Unge, David Yu, Jonas Sundén-Cullberg, Kristoffer Strålin

## Abstract

**Background:** In patients with infections, the Sequential Organ Failure Assessment (SOFA) score should be used to diagnose sepsis. However, manual SOFA calculation is time consuming. Thus, we developed an automated real-time SOFA score application that has been fully implemented into the major electronic health record (EHR) system (TakeCare) in Stockholm. We aimed to describe the method and evaluate its performance for detection of sepsis.

**Method:** We developed an automated SOFA application that presents a total score (SOFATotal) over time and a baseline score (SOFAChronic) based on outpatient data prior to hospital admission. We evaluated its performance on 583 sepsis alert patients in an emergency department, including 472 with sepsis according to manually calculated SOFA (SOFAManual). SOFATotal at 30 minutes and 3 hours, and quick-SOFA (qSOFA) were compared with SOFAManual for detection of sepsis. The acute SOFA score (SOFAAcute) was calculated by subtracting SOFAChronic from SOFATotal.

**Results:** SOFATotal at 3 hours showed moderate-good agreement with SOFAManual (R² = 0.76). Areas under receiver operating characteristic curves for sepsis detection were 0.83 at 30 minutes, 0.94 at 3 hours for SOFATotal, and 0.67 for qSOFA. Among patients with SOFAChronic ≥1 (21% of cases), SOFATotal of ≥2 was observed in 90% of cases (mortality 13.5%), although SOFAAcute of ≥2 was noted in 63% (mortality 16.7%).

**Conclusion:** The automated SOFA score demonstrated effectiveness in early sepsis detection, outperforming qSOFA, but adjustments for chronic baseline scores were necessary to avoid over-diagnosis. Further studies on broader patient populations would be useful to validate its clinical use.

## Introduction

The World Health Organization (WHO) recognised sepsis as a global health priority in 2017. In their sepsis resolution, they recommended the consistent use of the term “sepsis” and improvements in sepsis coding (1). Enhancing the International Classification of Diseases (ICD) coding for sepsis is crucial to provide accurate data on the burden of sepsis within a country. In Sweden, the burden of sepsis is under-reported, as correct ICD sepsis coding has been reported to occur in < 20% of patients who meet the criteria for sepsis (2).

Sepsis is operationally defined as organ dysfunction with a sequential organ failure assessment (SOFA) score increase of ≥2 due to an infection (3). Accordingly, it is recommended that SOFA score should be routinely calculated in hospitalised patients with infections, for adequate detection and diagnosis of sepsis.

A key challenge in utilising the SOFA score lies in its time-consuming calculation process. However, since SOFA score parameters are frequently recorded in the electronic health records (EHR), it is possible to use EHR data to calculate an automated SOFA score, as previously shown (2, 4–7). These studies used extracted data to calculate SOFA scores retrospectively. We are not aware of any previous real-time automated SOFA calculators using EHR data in clinical practice outside of the intensive care unit (ICU).

Another challenge for the use of SOFA is to distinguish chronic from acute organ failure. Gadrey et al. suggested that an acutely developed SOFA score of ≥2 is a poorer predictor of mortality than a total SOFA score of ≥2 (including both acute and chronic organ failure) for hospitalised patients with infection (8). Similarly, Christensen et al. proposed that the total SOFA score best predicts sepsis mortality and that chronic and acute organ failure are equally strongly associated with mortality (9). However, although chronic organ dysfunction is indicative for risk of mortality, it does not represent the biological process of sepsis, which may be responsive to specific sepsis treatments. Accordingly, for clinical trials of new drugs for sepsis, it is important to identify acute organ failure caused by an infection.

In an innovative project aimed to enable simple and accurate detection and diagnosis of sepsis, our group has developed a real-time electronic SOFA score application within the TakeCare EHR system, in collaboration with the CompuGroup Medica (CGM), the provider of TakeCare. TakeCare is the major EHR system of the Stockholm healthcare region. This electronic SOFA application was fully implemented in the TakeCare EHR system in five of six emergency hospitals in the Stockholm region, Sweden, on February 7, 2022. Since then, the SOFA score is automatically calculated and presented in the EHR of all hospitalised patients at these hospitals. In addition, when the EHR is opened for a patient visit in the period prior to 2022, the EHR system also provides electronic SOFA scores. To enable adjustment for individual baseline organ dysfunction, the system presents a chronic SOFA score, based on parameters from outpatient consultations prior to hospital admission.

The aims of the present study were to describe the design of the electronic SOFA application and evaluate its performance for detecting sepsis early after arrival in the emergency department (ED), including a comparison with quick-SOFA (qSOFA).

## Methods

### Design of the electronic SOFA application

The parameters of the electronic SOFA application are presented in Table 1. For coagulation, liver and renal dysfunction, the original SOFA score parameters are used (3, 13). However, for renal dysfunction, urine production is not included, since it is not consistently recorded in the EHR system. Furthermore, the SOFA application is not designed to access data concerning invasive methods of ventilatory support or the use of vasopressors. Accordingly, respiratory dysfunction could yield a maximum SOFA score of 3, and cardiovascular dysfunction could yield a maximum SOFA score of 1. For respiratory dysfunction, PaO_2_/FiO_2_is calculated when PaO_2_ is available. Supplementary Table S1 shows the table used by the system to estimate FiO_2_ based on registered oxygen delivery. In addition to PaO_2_/FiO_2_, pathologic oxygen saturation yields a respiratory SOFA score, based on previous study from our institution (14). The E-SOFA CNS score is calculated based on data from several surveillance tools fed into the TakeCare EHR system: Glasgow Coma Scale (GCS), Adapt, RETTS, and NEWS2.

**Table 1.**
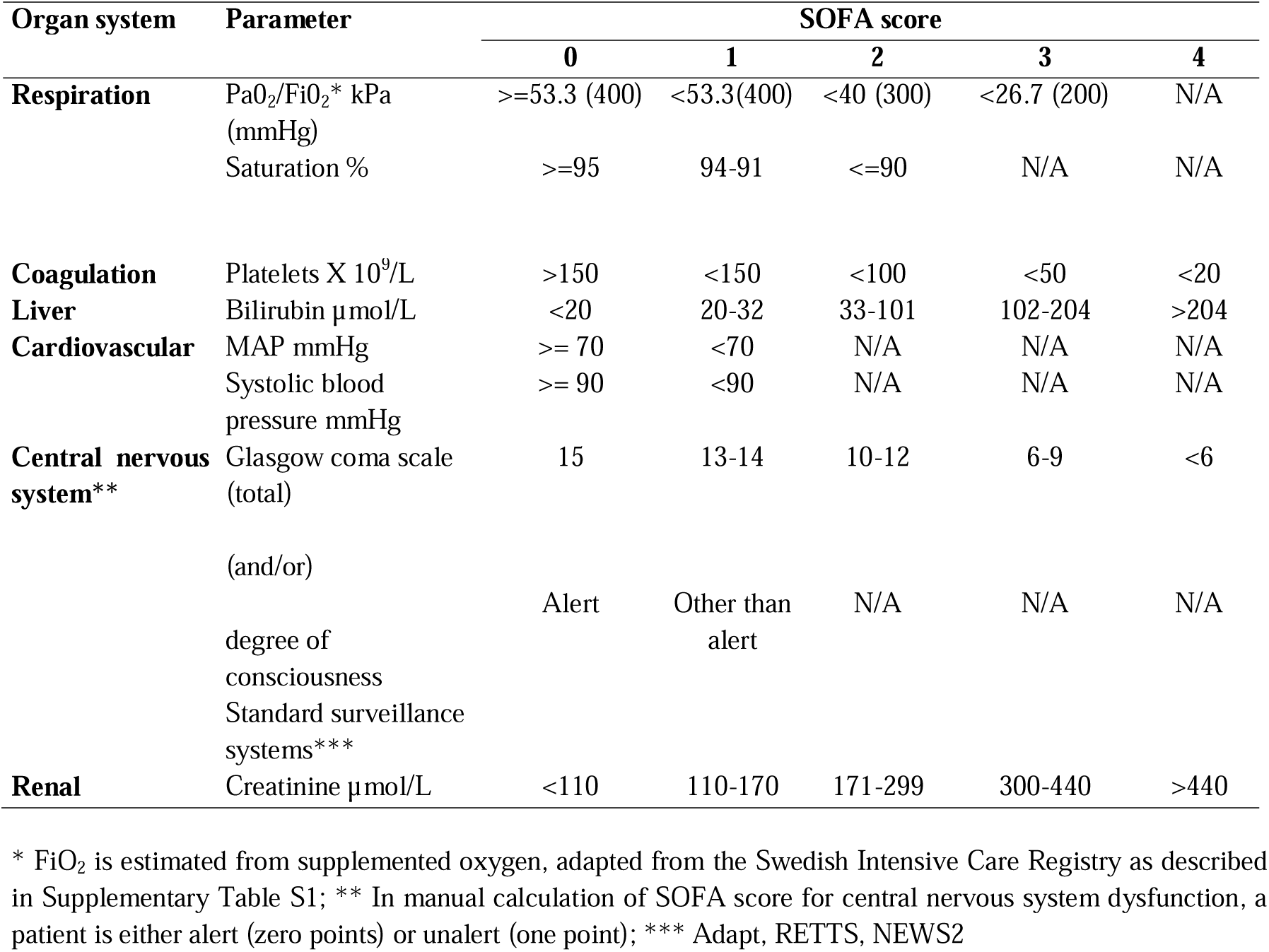
Criteria for manual calculation of SOFA score and automatic calculation of SOFA score in the electronic SOFA application.

| Organ system | Parameter | SOFA score |  |  |  |  |
| --- | --- | --- | --- | --- | --- | --- |
|  |  | 0 | 1 | 2 | 3 | 4 |
| **Respiration** | PaO<sub>2</sub>/FiO<sub>2</sub>* kPa (mmHg) | ≥53.3 (400) | <53.3(400) | <40 (300) | <26.7 (200) | N/A |
|  | Saturation % | ≥95 | 94-91 | ≤90 | N/A | N/A |
| **Coagulation** | Platelets X 10<sup>9</sup>/L | >150 | <150 | <100 | <50 | <20 |
| **Liver** | Bilirubin μmol/L | <20 | 20-32 | 33-101 | 102-204 | >204 |
| **Cardiovascular** | MAP mmHg | ≥ 70 | <70 | N/A | N/A | N/A |
|  | Systolic blood pressure mmHg | ≥ 90 | <90 | N/A | N/A | N/A |
| **Central nervous system\*\*** | Glasgow coma scale (total) | 15 | 13-14 | 10-12 | 6-9 | <6 |
|  | (and/or) |  |  |  |  |  |
|  | degree of consciousness Standard surveillance systems*** | Alert | Other than alert | N/A | N/A | N/A |
| **Renal** | Creatinine μmol/L | <110 | 110-170 | 171-299 | 300-440 | >440 |

\* FiO<sub>2</sub> is estimated from supplemented oxygen, adapted from the Swedish Intensive Care Registry as described in Supplementary Table S1; \*\* In manual calculation of SOFA score for central nervous system dysfunction, a patient is either alert (zero points) or unalert (one point); \*\*\* Adapt, RETTS, NEWS2

We designed the electronic SOFA application to automatically calculate SOFA scores based on routinely registered parameters in the EHR system, visualised in a SOFA score sheet (see an illustration in Supplementary Figure S1). Table 2 describes SOFA score terms of the electronic SOFA application. The definitions are similar to those proposed by Christensen et al. (9). The total SOFA score (SOFATotal) is calculated using the most pathological parameters that have been registered during the past 24 hours. For each new time point with a SOFA parameter registered, a new SOFATotal calculation is performed, based on the parameters from the new 24-hour window (Supplementary Figure S1). However, for creatinine, bilirubin, and platelet count, a registered parameter is applied in the SOFA calculations for 72 hours if no new registered parameter is added.

**Table 2.**
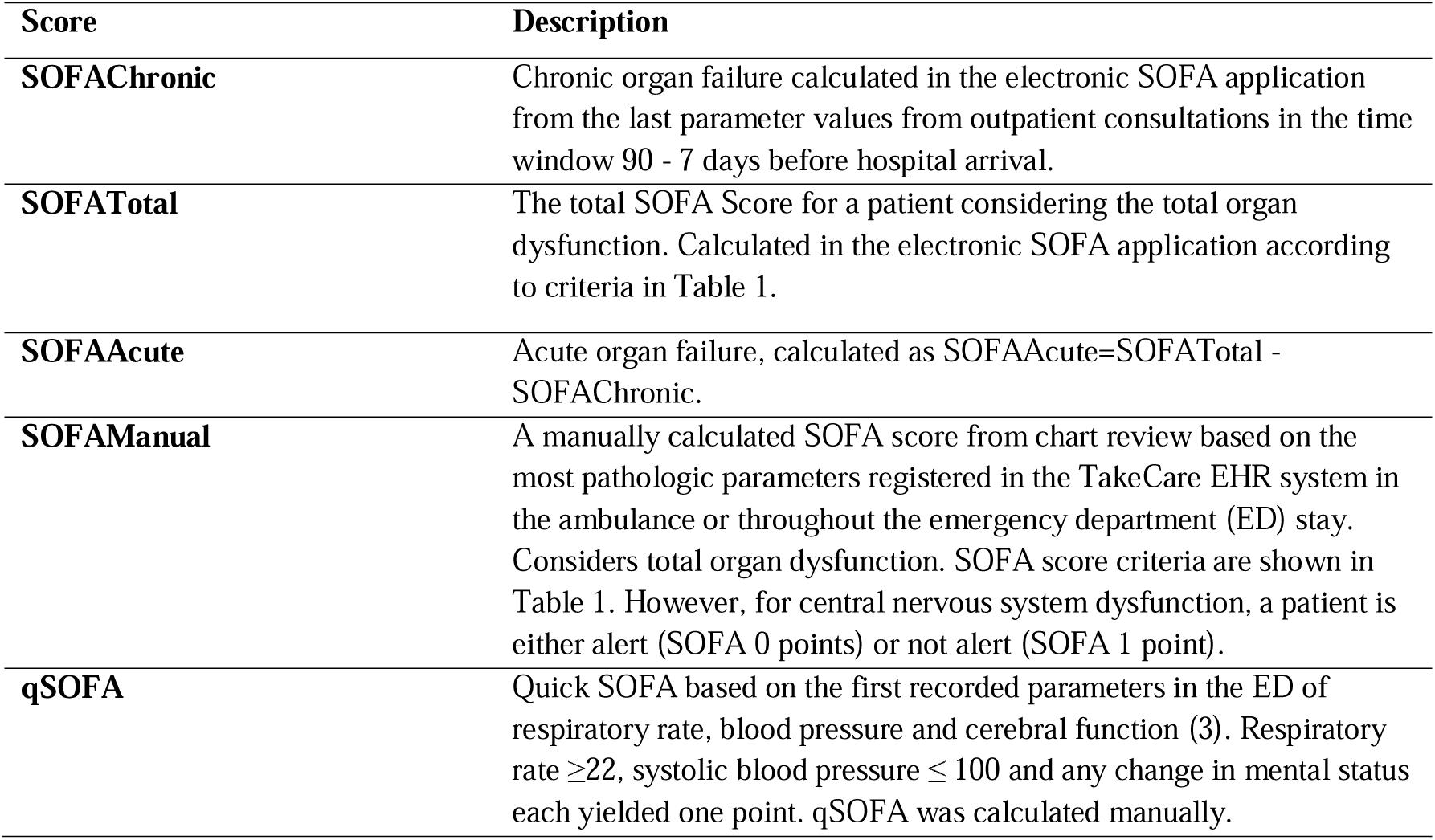
SOFA score terms used in this article.

A chronic SOFA score (SOFAChronic) is calculated using the most recent parameter values from outpatient consultations registered in a time window of 7-90 days before the current ED admittance. The system excludes parameters from inpatient care or ED visits and from 7 days prior to such events in the calculations of SOFAChronic.

Missing parameters are assumed to be normal for both SOFATotal and SOFAChronic.

### Study design and population of the evaluation study

In order to evaluate the performance of the electronic SOFA application, we performed a cohort study on patients who triggered a sepsis alert at the ED of Karolinska University Hospital Huddinge between September 2017 and March 2019 (Figure 1) (10). We chose to study this patient group, as these patients are rapidly assessed in the ED with routine collection of blood gas, including creatinine analysis, and routine collection of blood samples for platelet count and bilirubin. For the study population (Figure 1), we focused on patients with assumed bacterial infection according to the definition by the Centers for Disease Control and Prevention (11, 12), i.e. blood culture collected and antibiotic therapy provided for four days or until ICU admission or discharge, or death.

**Figure 1.**
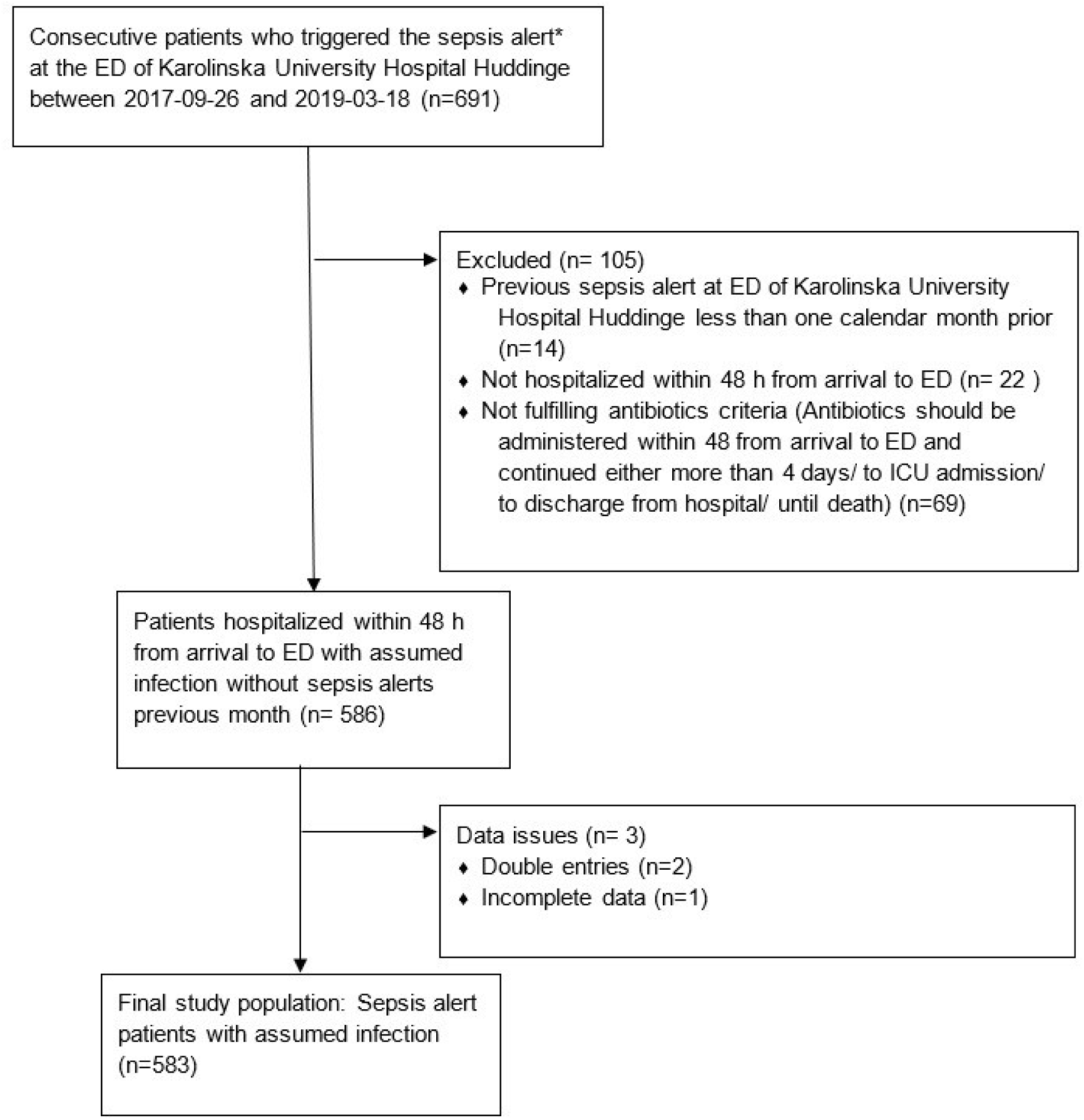
Flow chart of the study population at the emergency department (ED). * The sepsis alert was triggered for patients with signs or symptoms of ongoing infection (e.g., fever or clinical suspicion of infection) that were triaged according to the highest priority alone, or second/third highest priority combined with a plasma lactate exceeding 3.2 mmol/L

Patient data and clinical parameters were manually collected from the EHR system and have been used in previous studies (10, 15). SOFAManual was calculated based on the manual chart review, in which the most pathologic SOFA score parameters for each organ system during the ED stay had been registered (Table 2). The criteria for ManualSOFA score calculation are shown in Table 1. Pathologic parameters prior to the ED visit were not considered in the calculations of SOFAManual. qSOFA was calculated based on first recorded values in the ED of respiratory rate, systolic blood pressure and cerebral function (Table 2) (3).

The electronic SOFA scores are automatically presented when the TakeCare EHR system is accessed for a patient. For the present study, the EHR system was accessed to review each study patient, and the electronic SOFATotal scores at 30 min, 3 hours, and 24 hours, as well as SOFAChronic scores were recorded.

SOFA score data was double-checked in approximately 15% of the study patients to ensure data quality.

For the present study, an acute SOFA score (SOFAAcute) was calculated by subtracting SOFAChronic from SOFATotal.

### Statistics

Statistical analyses were performed using RStudio (version 2022.07.1). Proportions were compared using the Chi-square test. Agreement between SOFATotal3h and SOFAManual was illustrated with a heatmap, and the R² value was calculated. Sensitivity, specificity, predictive values, and area under the receiver operating characteristic curve (AUROC) were calculated for the detection of sepsis (using SOFAManual ≥2 as the reference standard) and 28-day mortality. Confidence intervals (CIs) were computed for diagnostic accuracy metrics. A p value of < 0.05 was considered statistically significant.

### Ethics

The study was approved by the Regional Ethical Committee of Stockholm (reference number 2017/1358-31). Since this was a retrospective study of patients triggering the sepsis alert in the ED, the ethical committee approved inclusion of all patients, consent to participate was not required.

## Results

### Study population of the evaluation study

A total of 583 sepsis alert patients met the study criteria and were included in the study population. The demographic information and clinical variables of the patients in the study cohort are summarised in Table 3. The median age was 74 years and 355 patients (60.9%) were male. The most common source of infection was the lower respiratory tract (34.8%). Manually calculated SOFA of ≥2 was noted in 472 patients (81.0%). For 172 patients (29.5%), do not attempt resuscitation (DNAR) orders were issued within two calendar days from admission to the ED.

**Table 3.** Characteristics and clinical variables of the study population (sepsis alert patients with assumed infection)

| Characteristics | Study population,<br>n = 583 |
| --- | --- |
| Median age (IQR) | 74 (62.5-82) |
| Sex, n (%) |  |
| Male | 355 (60.9) |
| Alcohol abuse & dependence*, n (%) | 37 (6.4) |
| Smoker, n (%)** | 193 (33.1) |
| Comorbidities, n (%) |  |
| Myocardial infarction | 79 (13.6) |
| Congestive heart failure | 110 (18.9) |
| Peripheral vascular disease | 59 (10.1) |
| Dementia | 80 (13.7) |
| Chronic lung disease*** | 95 (16.3) |
| Inflammatory systemic disease | 39 (6.7) |
| Stroke or TIA | 117 (20.1) |
| Kidney disease (moderate to severe) | 149 (25.6) |
| Diabetes | 166 (28.5) |
| Cancer | 131 (22.5) |
| Liver disease | 32 (5.5) |
| HIV positive | 1 (0.2) |
| Charlson comorbidity index (19) |  |
| Median (IQR) | 2 (1-4) |
| Mean | 2.8 |
| Source of infection, n (%) |  |
| Upper respiratory infection | 4 (0.7) |
| Lower respiratory infection | 203 (34.8) |
| Urinary Tract infection | 119 (20.4) |
| Intrabdominal infection | 48 (8.2) |
| Skin/soft tissue/skeletal/joint | 38 (6.5) |
| Central nervous system infection | 4 (0.7) |
| Heart | 12 (2.1) |
| Two or more infections | 13 (2.2) |
| Other**** | 12 (2.1) |
| Unknown | 130 (22.3) |
| Median hours in ED(IQR) | 4.9 (3.2-7.1) |
| Manually calculated SOFA $\geq 2$, n (%) | 472 (81.0) |
| Do not resuscitate orders within two days | 172 (29.5) |
| Patients two days after arrival at the ED |  |
| At hospital | 509 (87.3) |
| Discharged from hospital/to geriatric ward | 64 (11.0) |
| Dead | 10 (1.7) |
| Outcomes, n (%) |  |
| 28-day mortality | 81 (13.9) |
| Admission to ICU | 65 (11.3) |
| Blood culture, n (%) |  |
| Negative | 363 (62.3) |
| Positive |  |
| Gram-negative | 110 (18.9) |
| Gram-positive | 63 (10.8) |
| Mixed gram-neg. and gram-pos***** | 26 (4.5) |
| Fungi | 1 (0.2) |
| No blood culture collected | 20 (3.4) |

\* According to a diagnosis in a medical journal or a note from the emergency department (ED); \*\* Current and former smokers; \*\*\* Excluding mild asthma; \*\*\*\* Includes PICCLINE, tooth, Malaria, etc; \*\*\*\*\* One patient had mixed gram-positive and gram-negative species combined with Candida species

### SOFATotal - availability and distribution

SOFATotal increased over the first 24 hours with the most pronounced increase between 30 min and 3 hours (Supplementary Figure S2). The distribution of SOFATotal24h was similar to that of SOFAManual.

At 30 min, 97.1% of patients had at least one parameter recorded in SOFATotal30min, with an average parameter availability of 78% (Figure 2C, Table 4). Within SOFATotal30min, respiratory and renal SOFA scores were the most frequently recorded scores (Figure 2D).

**Figure 2.**
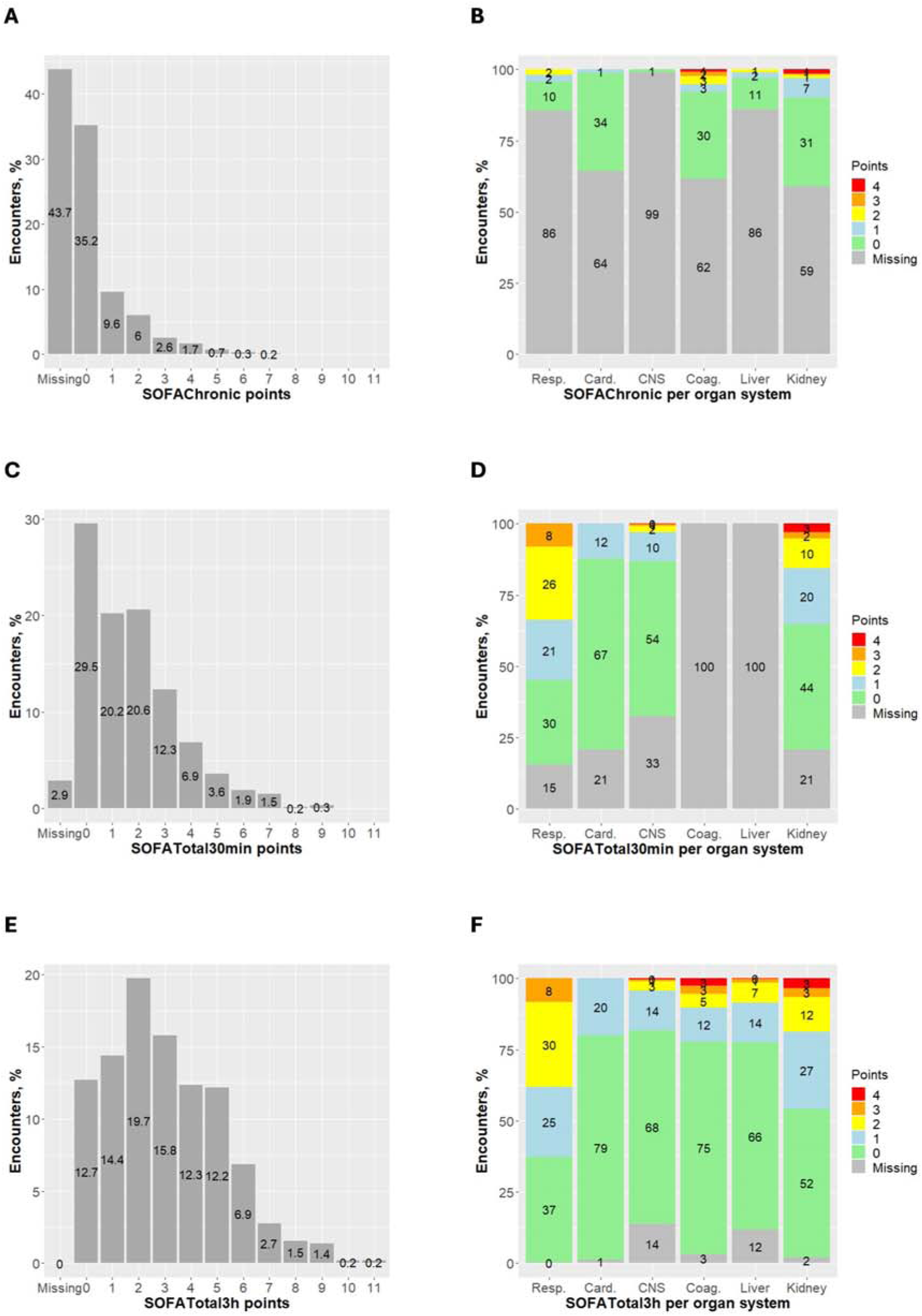
Distribution of SOFA points among sepsis alert patients with assumed infection. A) SOFAChronic, B) SOFAChronic per organ system, C) SOFATotal30min, D) SOFATotal30min per organ system, E) SOFATotal3h, and F) SOFATotal3h per organ system

**Table 4.**
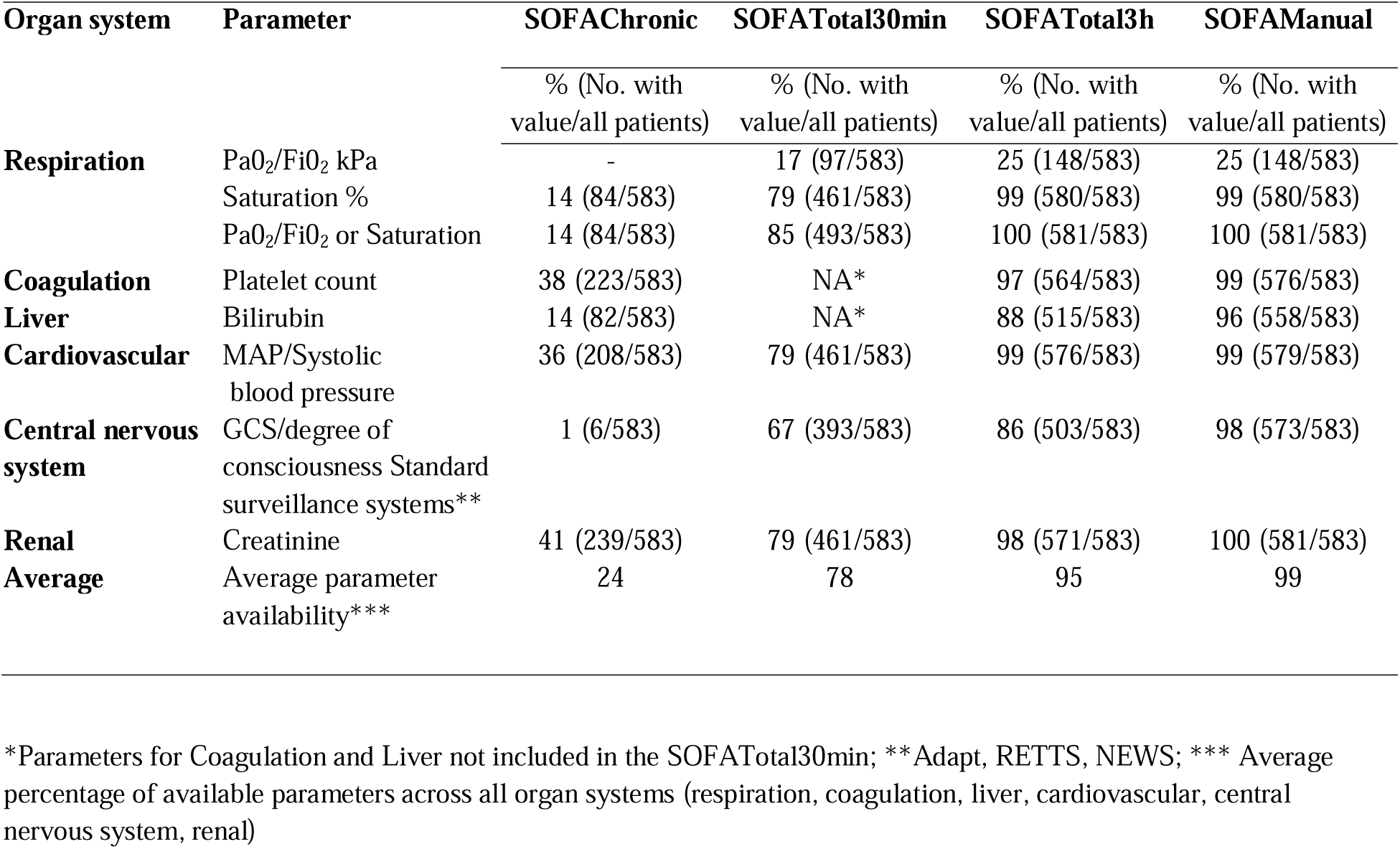
Parameter availability of SOFA scores per organ system for sepsis alert patients with assumed infection.

By 3 hours, all patients had at least one recorded value in SOFATotal3h, with an average availability of 95% (Figure 2E, Table 4). Respiratory and renal SOFA scores remained the most prevalent scores (Figure 2F).

The difference in median scores between SOFATotal30min and SOFATotal3h was primarily due to the inclusion of coagulation and liver parameters in SOFATotal3h. Liver and coagulation scores were recorded in 22% of patients (Figure 2F). Additionally, the proportions of patients with respiratory, cardiovascular, CNS, and renal SOFA scores increased by 15%, 8%, 5%, and 11%, respectively, between the two time points.

### Association between SOFATotal at 3 hours and SOFAManual

SOFATotal3h and SOFAManual showed a moderate correlation, with 56% of patients (325 out of 583) having identical scores (Figure 3). The agreement between SOFATotal3h and SOFAManual was moderate-good with an R^2^ value of 0.76 (p<0.001). Meanwhile the agreement between SOFATotal24h and SOFAManual was slightly better (R^2^ 0.81; p<0.001). For SOFATotal3h and SOFAManual, the following R^2^ values for organ systems were noted: respiration 0.68 (p<0.001); cardiovascular 0.50 (p<0.001); CNS 0.29 (p<0.001); coagulation 0.88 (p<0.001); liver 0.61 (p<0.001); renal 0.93 (p<0.001). CNS SOFA scores was more prevalent using SOFAManual (25 %, 143 patients each with one point) than SOFATotal3h (18%, 106 patients with scores ranging from one to four points).

**Figure 3.**
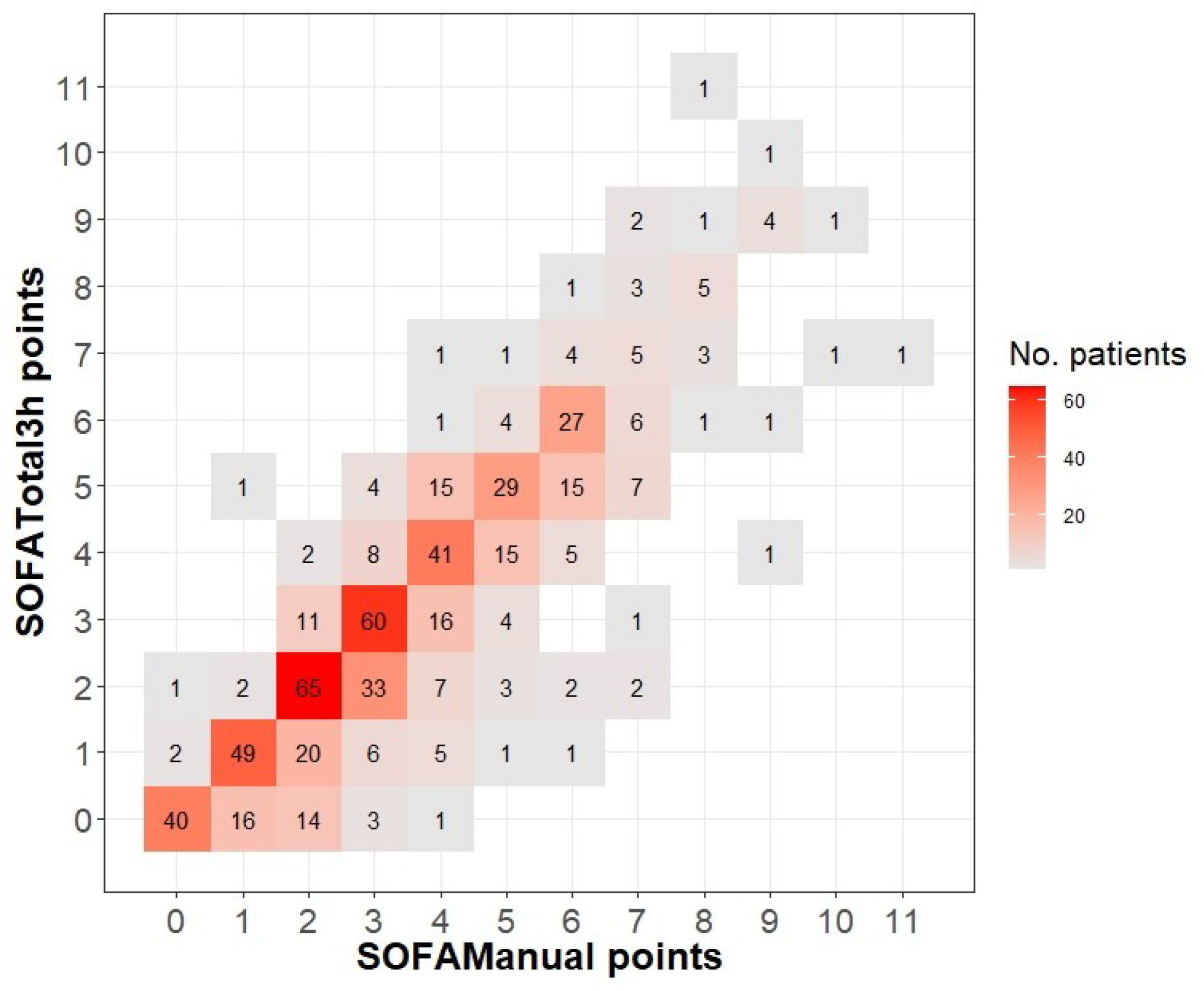
Heatmap showing agreement between SOFATotal3h and SOFAManual for sepsis alert patients with assumed infection (R^2^ value = 0.76, p<0.001)

### SOFAChronic - availability and distribution

SOFAChronic was 0 in 35%, ≥1 in 21%, and missing in 44% of patients. Supplementary Table S2 outlines the characteristics of patients with different SOFAChronic scores. Notably, patients with missing SOFAChronic had similar age and sex profiles compared to those with SOFAChronic 0 or ≥1. However, they exhibited a significantly higher incidence of dementia (22%; 55/255 vs 7.6%; 25/328; p<0.01) and a higher proportion of DNAR orders (35%; 90/255 vs 25%; 82/328; p<0.01).

At least one SOFAChronic parameter was recorded for 328 patients (56.3%), with an average parameter availability of 24% (Figure 2A, Table 4). Among the organ systems, parameter availability was highest for renal (41%), followed by coagulation (38%) and cardiovascular (36%) systems. Conversely, the CNS system had the lowest availability at 1% (Figure 2B, Table 4). The most common forms of organ dysfunction were renal dysfunction (57 patients, 9.8%) and coagulation dysfunction (46 patients, 7.9%) (Figure 2B).

### Distribution and mortality of total and acute SOFA in relation to chronic SOFA

Figure 4 shows the distributions of SOFATotal3h and SOFAAcute3 and Figure 5 shows mortalities at ≥2 points of SOFATotal3h and SOFAAcute3h, at different SOFAChronic scores. Among patients with SOFAChronic missing, 0, and ≥1, a SOFATotal3h of ≥2 was observed in 69%, 67%, and 90%, with mortality rates of 19.8%, 16.1%, and 13.5%, respectively. In patients with SOFAChronic ≥1, SOFAAcute ≥2 was noted in 63% (mortality 16.7%: Figures 4, 5).

**Figure 4.**
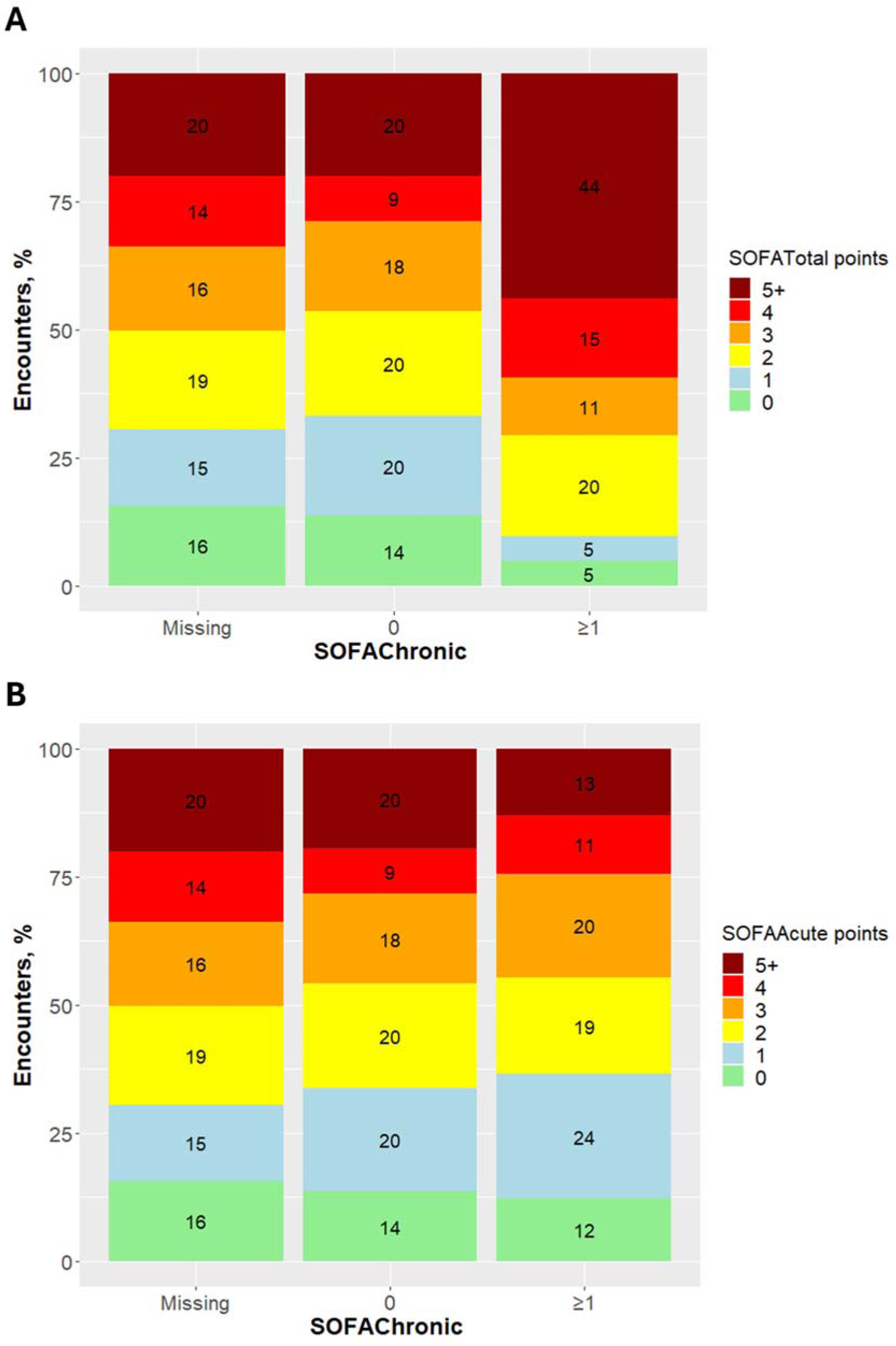
Distribution of A) SOFATotal3h points and B) SOFAAcute3h points at different SOFAChronic scores, in sepsis alert patients with assumed infection

**Figure 5.**
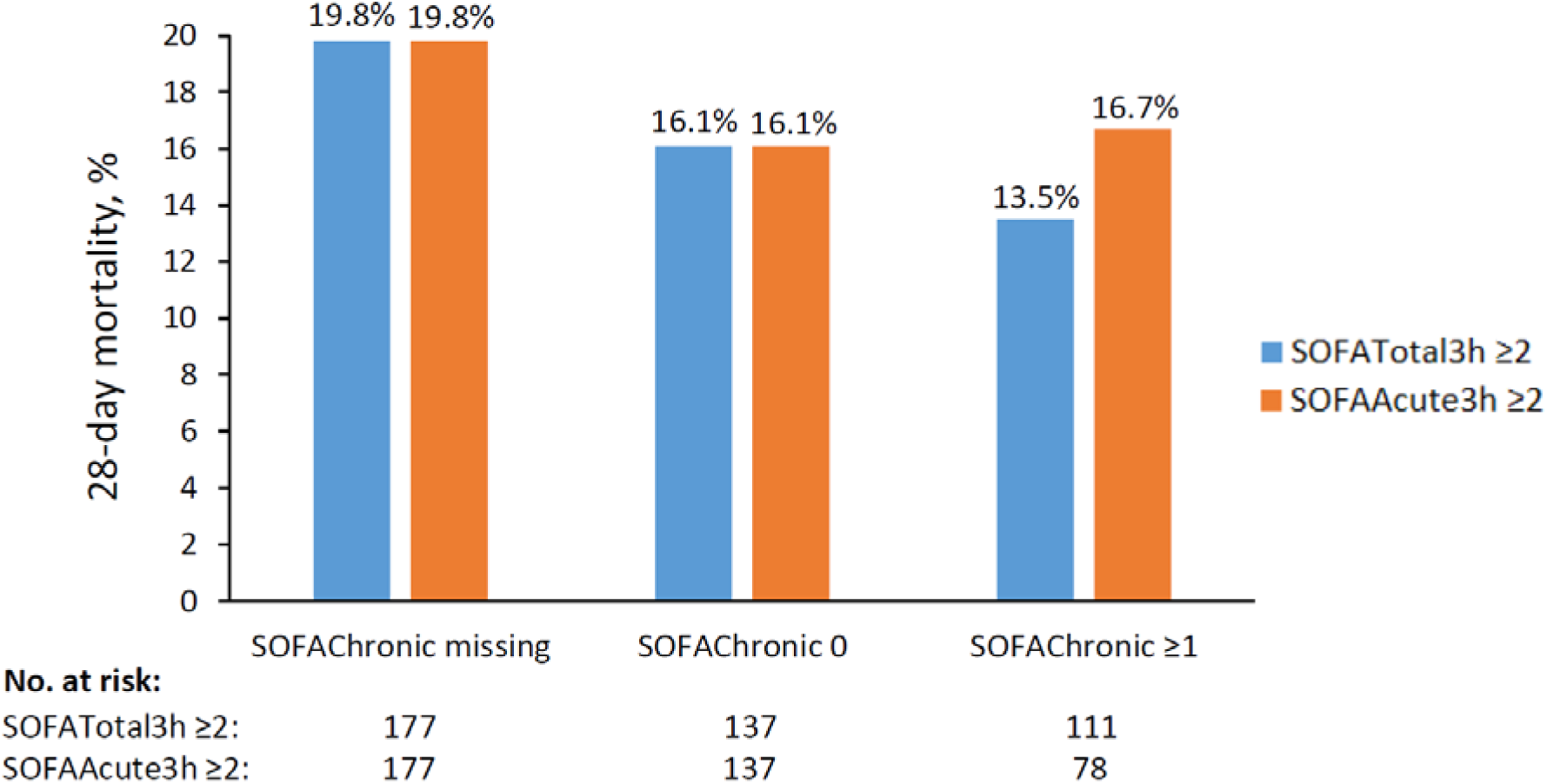
28-day mortalities at ≥2 points of SOFATotal3h and SOFAAcute3h, at different SOFAChronic scores, in sepsis alert patients with assumed infection

### Detection of sepsis

For the detection of sepsis with SOFAManual ≥ 2 used as reference standard, the sensitivities were 89% for SOFATotal3h, 58% for SOFATotal30min, and 27% for qSOFA (Table 5). However, SOFATotal3h, SOFATotal30min, and qSOFA showed similar specificities of 95-97%. In addition, for detection of sepsis, the following AUROCs were noted: SOFATotal3h 0.94; SOFATotal30min 0.88; and qSOFA 0.67 (Figure 6).

**Figure 6.**
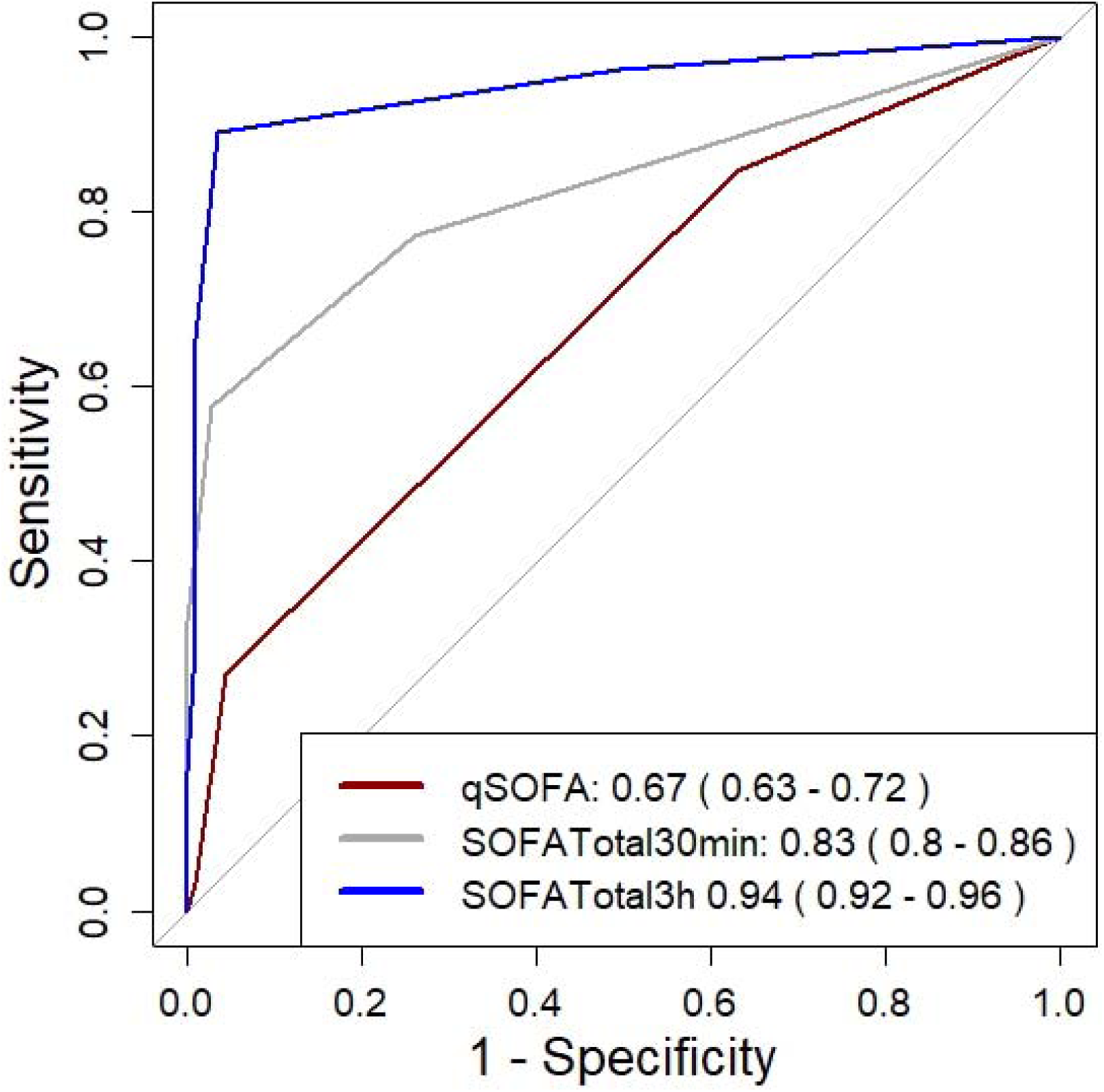
Receiver operating characteristic (ROC) for the detection of sepsis with SOFAManual as reference standard. The legend indicates the corresponding area under ROC curve (AUROC) values with 95% confidence intervals

**Table 5.** Predictive performance of qSOFA and SOFATotal for detecting sepsis, with SOFAManual ≥2 as reference standard, in sepsis alert patients with assumed infection.

| Score, points | Sensitivity<br>% (ratio)* | Specificity<br>% (ratio)** | PPV<br>% (ratio)*** | NPV<br>% (ratio)**** |
| --- | --- | --- | --- | --- |
| **qSOFA $\geq 2$ points** | 27 (128/472) | 95 (106/111) | 96 (128/133) | 24 (106/450) |
| **SOFATotal30 min $\geq 2$ points** | 58 (273/472) | 97 (108/111) | 99 (273/276) | 35 (108/307) |
| **SOFATotal3h $\geq 2$ points** | 89 (421/472) | 96 (107/111) | 99 (421/425) | 68 (107/158) |

Predictive performance of qSOFA, SOFATotal30min and SOFATotal3h h using an elevation of 2 or more points in SOFAManual and SOFATotal24h as test gold standards for organ dysfunction. \* Sensitivity=True positive/(True positive + False negative); \*\* Specificity=True negative/(True negative + False positive); \*\*\* Positive Predictive Value =True positive/(True positive + False positive); \*\*\*\*Negative Predictive Value=True negative/(True negative + False negative). Reported as percentage, numerator and denominator within brackets

Using the automated SOFA application, a SOFA score of ≥ 2 was noted in 425 patients (72.9%) with SOFATotal3h and 392 patients (67.2%) with SOFAAcute3h.

### Prediction of 28-day mortality

For prediction of 28-day mortality, the AUROCs were similar (0.65-0.68) for SOFAManual, qSOFA, and SOFATotal at 30 min and 3 hours (Supplementary Figure S3A). In addition, the AUROC values for both SOFAAcute30min and SOFAAcute3h were 0.66. SOFAAcute30min ≥2 showed a sensitivity of 65% and a specificity of 62%, although qSOFA ≥2 showed a sensitivity of 46% and a specificity of 81% (Supplementary Table S3). Among patients with SOFAChronic ≥1, the AUROC was 0.71 for SOFATotal3h and 0.68 for SOFAAcute3h (Supplementary Figure S3B).

## Discussion

This article describes the design of an automated real-time SOFA score calculator that has been fully implemented into a major EHR system, and evaluates its performance for detection of sepsis. The automated SOFATotal score showed moderate-good agreement with manually calculated SOFA and was effective in detecting sepsis, as early as 30 min after ED arrival, with better performance than qSOFA. However, in patients with chronic organ dysfunction, SOFATotal tended to over-diagnose sepsis, illustrating the need to adjust for SOFAChronic parameters when available.

SOFATotal30min (based on cardiovascular, respiratory, CNS, and renal dysfunction) demonstrated higher sensitivity than qSOFA for detecting sepsis, while the specificities were comparable. ROC curve analysis confirmed the superiority of SOFATotal30min over qSOFA in detecting sepsis, although the AUROCs for predicting mortality were similar between the two scores. Previous studies have shown that manually calculated SOFA performs better than qSOFA (16, 17), but our study showed that automated SOFA score was superior to qSOFA already at 30 min, before liver or coagulation data were available. This indicates that automated SOFA score calculation early in the ED can be useful for detection of sepsis.

The SOFA score system was initially developed to quantify organ dysfunction in the ICU (13), and its use was extended to non-ICU patients for detection of sepsis within the Sepsis-3 classification (3). In the Sepsis-3 classification, a patient’s SOFA score should be compared with the baseline SOFA score to identify a SOFA score increase of ≥2 (18). However, it has not been defined how the baseline SOFA should be determined. In order to account for baseline SOFA, we designed SOFAChronic within our electronic SOFA application. In our evaluation study, among patients with a SOFAChronic score of ≥ 1, the adjustment for SOFAChronic reduced the proportion of patients with a SOFA score of ≥2 from 90% (SOFATotal) to 63% (SOFAAcute). In addition, the proportion with 28-day death among patients with a SOFA score of ≥2 increased from 13.5% (SOFATotal) to 16.7% (SOFAAcute). Thus, this adjustment made the distribution of SOFA scores and mortality rate at SOFA ≥2 more consistent with those of patients with a SOFAChronic score of 0 or missing. These findings suggest that over-diagnosis of sepsis using SOFATotal could be mitigated by using SOFAAcute in patients with SOFAChronic parameters available. Over-diagnosis of sepsis is concerning, since it may lead to over-use of both broad-spectrum antibiotics and advanced supervision.

Within our electronic SOFA application, low oxygen saturation is used to generate respiratory SOFA scores in addition to PaO_2_. This is motivated by our recent study on the use of oxygen saturation to improve detection of respiratory organ dysfunction in the SOFA score (14). Recently, an update of the SOFA score for defining sepsis has been proposed, which also suggests the use of oxygen saturation (19).

Since the Swedish government in 2019 selected sepsis as one of ten diagnoses for clinical pathways in healthcare, a national working group has developed a patient-centered clinical pathway for sepsis, which is currently being implemented (20). Accurate sepsis coding is a major objective within this clinical pathway, and to achieve this goal, it has been proposed that an electronic SOFA score calculator within the EHR system should be used (20). The present electronic SOFA score calculator has served as a prototype, and a detailed description of it has been sent to all healthcare regions in Sweden. In Region Stockholm hospitals, the electronic SOFA score application in the TakeCare EHR system has simplified sepsis coding, and sepsis coding has improved since the implementation of this system (data not shown).

The study has some strengths. First, the study was based on real-time data from a fully implemented electronic SOFA score application that is being used in clinical practice. Second, as the electronic SOFA application identifies sepsis as early as 30 min from ED arrival, it can be used for early clinical optimisation regarding antimicrobial therapy and supervision, and for targeting patients for clinical trials of new interventions and drugs for sepsis. Third, although the electronic SOFA score application was evaluated for detection of sepsis, it can be used to assess and monitor organ dysfunction from any cause, not just from infections.

The study has several limitations. First, the study was a single-site study based on sepsis alert patients. The electronic SOFA application should to be validated across broader patient populations of unselected patients. Second, many patients had missing SOFA parameters, as in the derivation and validation studies of the Sepsis-3 classification (18). Missing parameters were most pronounced for SOFAChronic scores. However, the patients with missing SOFAChronic scores had SOFATotal distributions similar to those of patients with SOFAChronic of 0. Third, the study showed that the automatic SOFA application under-diagnosed CNS dysfunction. If free-text information could be added to the electronic SOFA application, the CNS SOFA scores would be more reliable. Fourth, vasopressors, ventilatory support, and urine output were not included in the electronic SOFA application. However, we believe that these omissions had a limited effect on the SOFA scores of the present study, as we focused on the first hours after hospital arrival. Fifth, since SOFAManual assessed total organ dysfunction, independently on when the organ dysfunction started, performance of the electronic SOFA application for detection of sepsis (with SOFAManual as reference standard) could only be evaluated for SOFATotal, not for SOFAAcute.

## Conclusion

The study showed that the automated SOFATotal score calculation within the EHR system showed moderate-good agreement with SOFAManual and was effective in detecting sepsis, as early as 30 min after ED arrival, with better performance than qSOFA. When SOFAChronic was ≥1, SOFATotal tended to over-diagnose sepsis, illustrating the need to adjust for SOFAChronic parameters when available. The study supports use of automated SOFA score within the EHR system to diagnose sepsis, but additional studies on broader patient populations would be useful.

## Abbreviations

AUROC: Area under the receiver operating characteristic curve
CI: Confidence interval
DNAR: Do not attempt resuscitation
ED: Emergency department
EHR: Electronic health records
GCS: Glasgow coma scale
HIV: Human immunodeficiency virus
ICD: International classification of diseases
ICU: Intensive care unit
IQR: Interquartile range
MAP: Mean arterial pressure
NEWS: National early warning score
OR: Odds ratio
RETTS: Rapid emergency triage and treatment system
ROC: Receiver operating characteristic
SOFA: Sequential organ failure assessment
TIA: Transient ischemic attack
WHO: World health organization

## Acknowledgments

The authors thank Eva Saras for leading the implementation process of the electronic SOFA score calculator into the TakeCare EHR system and CompuGroup Medica (CGM) for collaboration regarding the design and implementation of the electronic SOFA score calculator.

## Financial support

The implementation of the electronic SOFA score calculator into the TakeCare EHR system was supported by Innovationsfonden (20181028), the Swedish Innovation Agency (VINNOVA) under the frame of NordForsk (90456), the Swedish Innovation Agency (VINNOVA; 2018–03350), and Region Stockholm.

## Conflicts of interest

All authors declare that they have no conflicts of interest.

## Availability of data and materials

All data produced in the present study are available upon reasonable request to the authors

## SUPPLEMENTARY MATERIALS

**Supplementary Figure S1.**
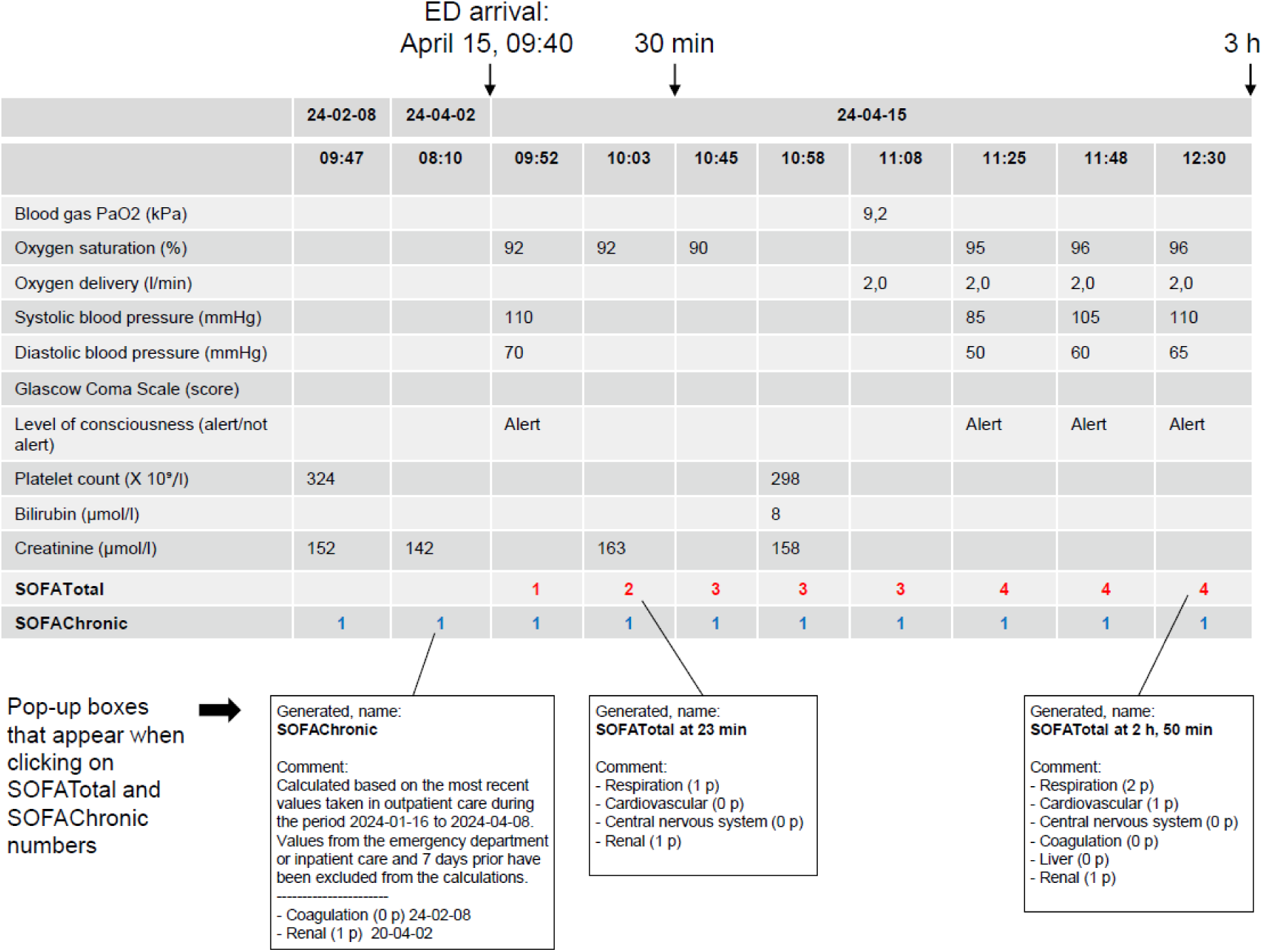
Illustration of the automated SOFA score application translated from Swedish to English, regarding a patient with fever who arrives at the emergency department (ED). Each column represents registered parameters per time of registration. SOFATotal is calculated based on the most pathological parameters that have been registered during the past 24 hours. SOFAChronic is calculated using the most recent parameters from outpatient consultations in a time window of 7-90 days before the current hospital admission

**Supplementary Figure S2.**
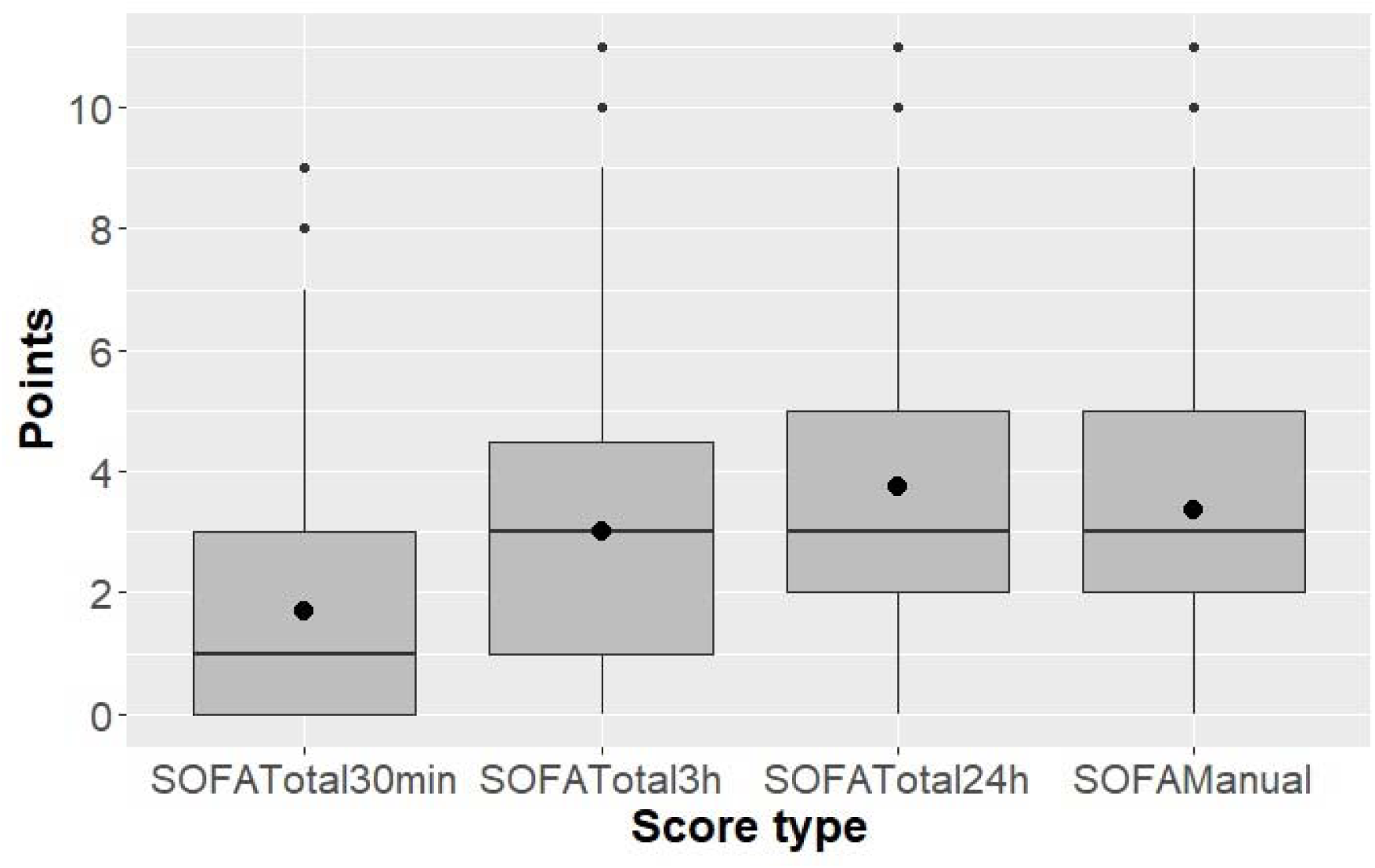
Box-plots showing the distribution of SOFA scores at different time points among sepsis alert patients with assumed infection. The filled black circles indicate the mean values

**Supplementary Figure S3.**
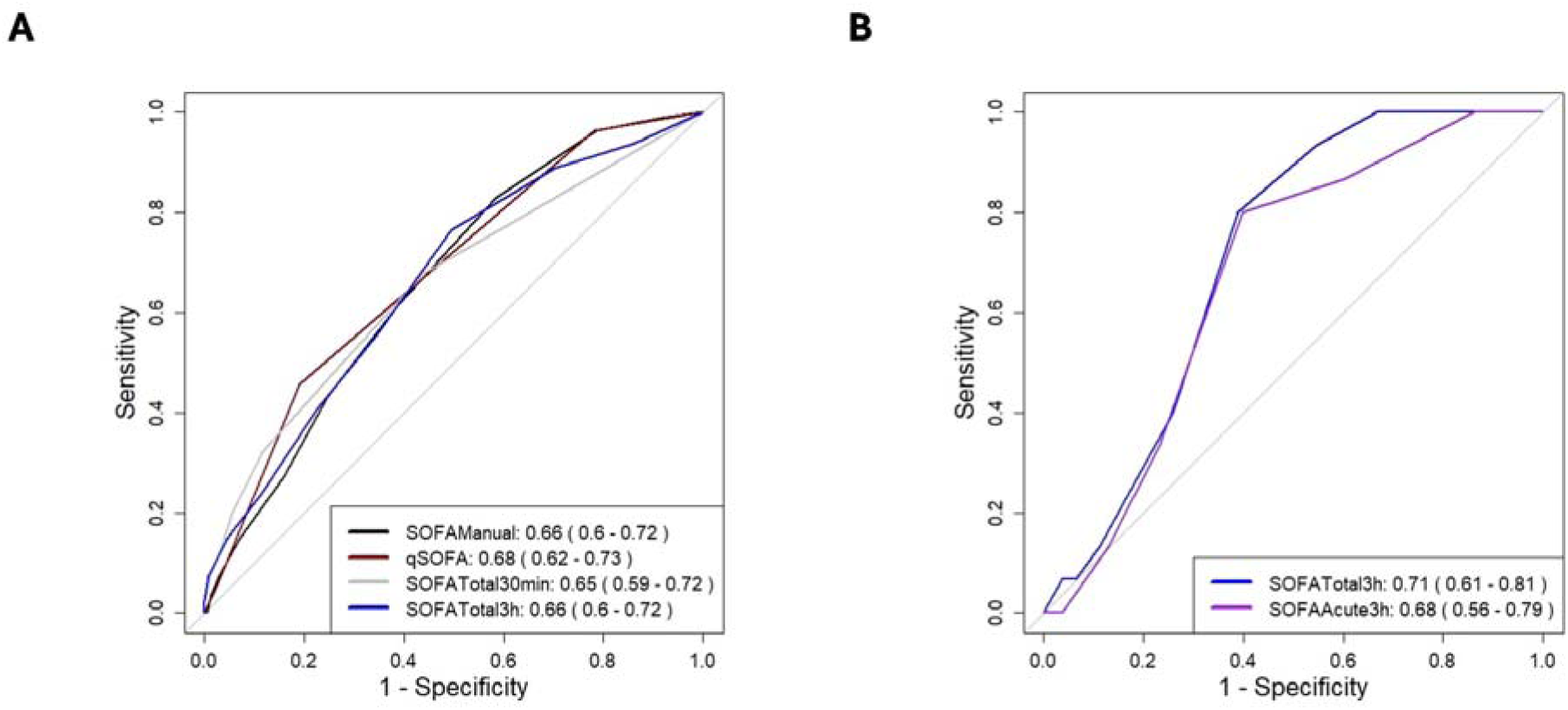
Receiver operating characteristic (ROC) curve for the prediction of 28-day mortality in sepsis alert patients with assumed infection. A) SOFAManual, qSOFA, and SOFATotal at 30 min and 3 hours on all study patients, B) SOFATotal and SOFAAcute at 3 hours on 123 patients with SOFAChronic of ≥1. The legend indicates the corresponding area under ROC curve (AUROC) values with 95% confidence intervals

**Supplementary Table S1.** Fraction of inspired oxygen (FI0_2_) within the automatic SOFA score calculator of the TakeCare electronic health records.

| Oxygen l/min | FI0<sub>2</sub> |
| --- | --- |
| **0** | 0.21 |
| **1** | 0.22 |
| **2** | 0.24 |
| **3** | 0.26 |
| **4** | 0.28 |
| **5** | 0.35 |
| **6** | 0.37 |
| **7** | 0.40 |
| **8 or more** | 0.45 |

Assumption on Fraction of inspired oxygen (FI0<sub>2</sub>) is based on liters of oxygen administered regardless of how oxygen is administered

**Supplementary Table S2.**
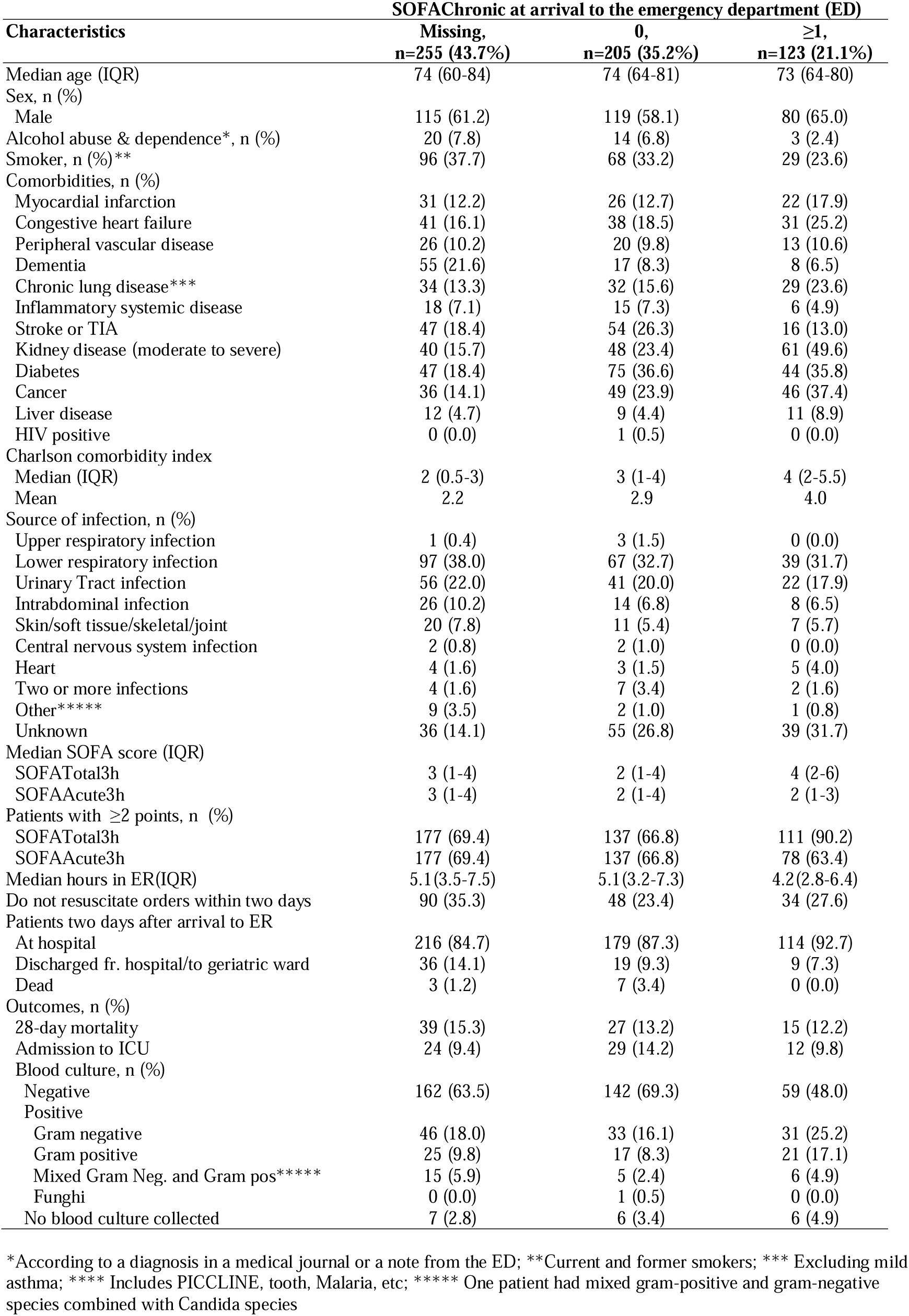
Characteristics and clinical variables of the study population (sepsis alert patients with assumed infection) stratified by SOFAChronic.

| Characteristics | SOFACHronic at arrival to the emergency department (ED) |  |  |
| --- | --- | --- | --- |
|  | Missing,<br>n=255 (43.7%) | 0,<br>n=205 (35.2%) | ≥1,<br>n=123 (21.1%) |
| Median age (IQR) | 74 (60-84) | 74 (64-81) | 73 (64-80) |
| Sex, n (%) |  |  |  |
| Male | 115 (61.2) | 119 (58.1) | 80 (65.0) |
| Alcohol abuse & dependence*, n (%) | 20 (7.8) | 14 (6.8) | 3 (2.4) |
| Smoker, n (%)** | 96 (37.7) | 68 (33.2) | 29 (23.6) |
| Comorbidities, n (%) |  |  |  |
| Myocardial infarction | 31 (12.2) | 26 (12.7) | 22 (17.9) |
| Congestive heart failure | 41 (16.1) | 38 (18.5) | 31 (25.2) |
| Peripheral vascular disease | 26 (10.2) | 20 (9.8) | 13 (10.6) |
| Dementia | 55 (21.6) | 17 (8.3) | 8 (6.5) |
| Chronic lung disease*** | 34 (13.3) | 32 (15.6) | 29 (23.6) |
| Inflammatory systemic disease | 18 (7.1) | 15 (7.3) | 6 (4.9) |
| Stroke or TIA | 47 (18.4) | 54 (26.3) | 16 (13.0) |
| Kidney disease (moderate to severe) | 40 (15.7) | 48 (23.4) | 61 (49.6) |
| Diabetes | 47 (18.4) | 75 (36.6) | 44 (35.8) |
| Cancer | 36 (14.1) | 49 (23.9) | 46 (37.4) |
| Liver disease | 12 (4.7) | 9 (4.4) | 11 (8.9) |
| HIV positive | 0 (0.0) | 1 (0.5) | 0 (0.0) |
| Charlson comorbidity index |  |  |  |
| Median (IQR) | 2 (0.5-3) | 3 (1-4) | 4 (2-5.5) |
| Mean | 2.2 | 2.9 | 4.0 |
| Source of infection, n (%) |  |  |  |
| Upper respiratory infection | 1 (0.4) | 3 (1.5) | 0 (0.0) |
| Lower respiratory infection | 97 (38.0) | 67 (32.7) | 39 (31.7) |
| Urinary Tract infection | 56 (22.0) | 41 (20.0) | 22 (17.9) |
| Intrabdominal infection | 26 (10.2) | 14 (6.8) | 8 (6.5) |
| Skin/soft tissue/skeletal/joint | 20 (7.8) | 11 (5.4) | 7 (5.7) |
| Central nervous system infection | 2 (0.8) | 2 (1.0) | 0 (0.0) |
| Heart | 4 (1.6) | 3 (1.5) | 5 (4.0) |
| Two or more infections | 4 (1.6) | 7 (3.4) | 2 (1.6) |
| Other***** | 9 (3.5) | 2 (1.0) | 1 (0.8) |
| Unknown | 36 (14.1) | 55 (26.8) | 39 (31.7) |
| Median SOFA score (IQR) |  |  |  |
| SOFA<sub>Total3h</sub> | 3 (1-4) | 2 (1-4) | 4 (2-6) |
| SOFA<sub>Acute3h</sub> | 3 (1-4) | 2 (1-4) | 2 (1-3) |
| Patients with ≥2 points, n (%) |  |  |  |
| SOFA<sub>Total3h</sub> | 177 (69.4) | 137 (66.8) | 111 (90.2) |
| SOFA<sub>Acute3h</sub> | 177 (69.4) | 137 (66.8) | 78 (63.4) |
| Median hours in ER(IQR) | 5.1(3.5-7.5) | 5.1(3.2-7.3) | 4.2(2.8-6.4) |
| Do not resuscitate orders within two days | 90 (35.3) | 48 (23.4) | 34 (27.6) |
| Patients two days after arrival to ER |  |  |  |
| At hospital | 216 (84.7) | 179 (87.3) | 114 (92.7) |
| Discharged fr. hospital/to geriatric ward | 36 (14.1) | 19 (9.3) | 9 (7.3) |
| Dead | 3 (1.2) | 7 (3.4) | 0 (0.0) |
| Outcomes, n (%) |  |  |  |
| 28-day mortality | 39 (15.3) | 27 (13.2) | 15 (12.2) |
| Admission to ICU | 24 (9.4) | 29 (14.2) | 12 (9.8) |
| Blood culture, n (%) |  |  |  |
| Negative | 162 (63.5) | 142 (69.3) | 59 (48.0) |
| Positive |  |  |  |
| Gram negative | 46 (18.0) | 33 (16.1) | 31 (25.2) |
| Gram positive | 25 (9.8) | 17 (8.3) | 21 (17.1) |
| Mixed Gram Neg. and Gram pos***** | 15 (5.9) | 5 (2.4) | 6 (4.9) |
| Funghi | 0 (0.0) | 1 (0.5) | 0 (0.0) |
| No blood culture collected | 7 (2.8) | 6 (3.4) | 6 (4.9) |

\*According to a diagnosis in a medical journal or a note from the ED; \*\*Current and former smokers; \*\*\* Excluding mild asthma; \*\*\*\* Includes PICCLINE, tooth, Malaria, etc; \*\*\*\*\* One patient had mixed gram-positive and gram-negative species combined with Candida species

**Supplementary Table S3.**
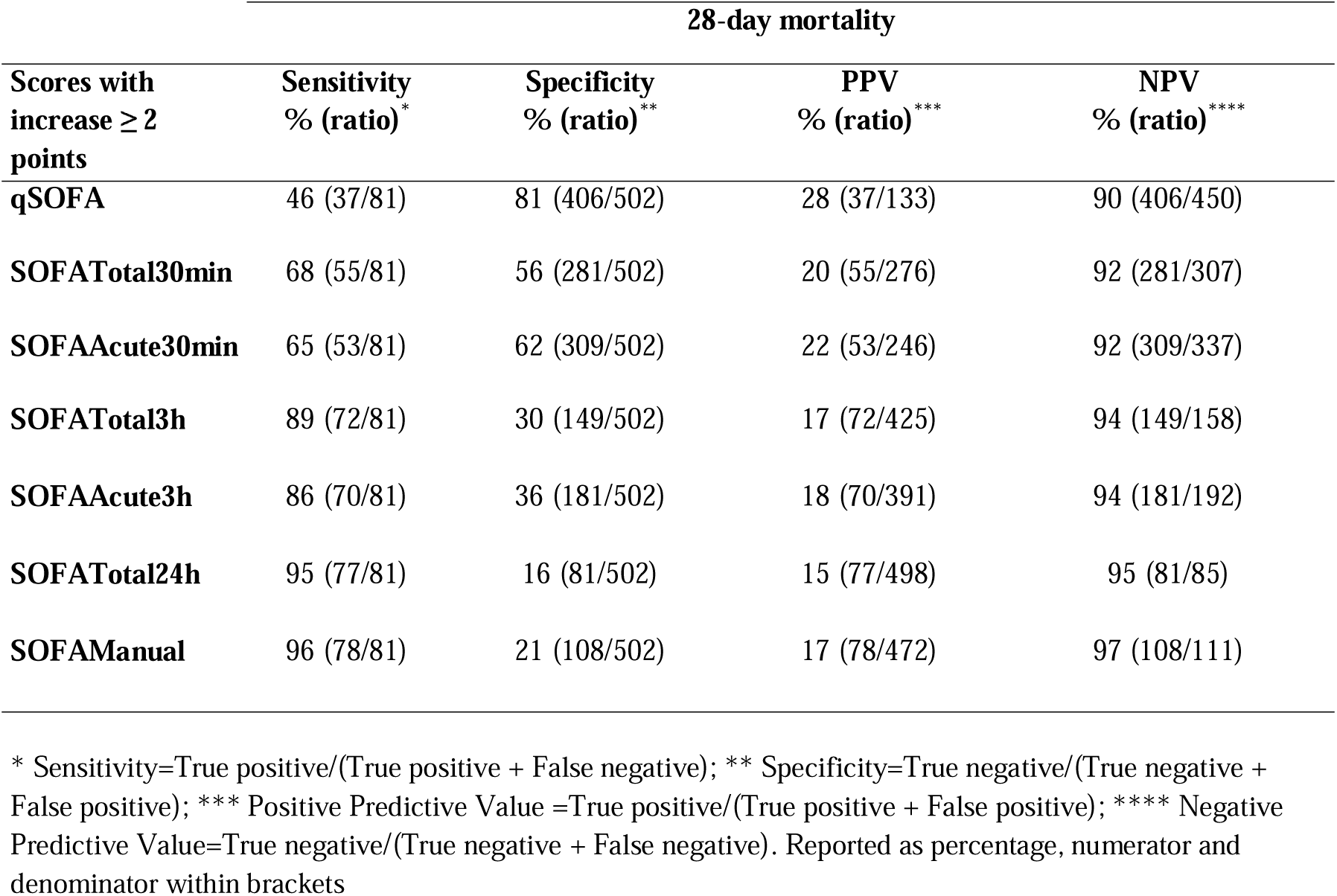
Predictive performance of qSOFA, SOFATotal, SOFAAcute and SOFAManual for 28-day mortality in sepsis alert patients with assumed infection.

## Notes

### Competing Interest Statement

The authors have declared no competing interest.

